# Hypertension Shifts Gut Microbiota and Tryptophan Metabolism in Women

**DOI:** 10.1101/2025.01.30.25321440

**Authors:** Shrushti Shah, Chunlong Mu, Grace Shen-Tu, Kristina Schlicht, Nils Forkert, Matthias Laudes, Harald Köfeler, Jane Shearer

**Affiliations:** Faculty of Kinesiology, University of Calgary, Calgary, AB. Canada; Libin Cardiovascular Institute, University of Calgary, Calgary, AB. Canada; Department of Biochemistry and Molecular Biology, Cumming School of Medicine, University of Calgary, Calgary, AB. Canada; Alberta Children’s Hospital Research Institute, University of Calgary, Calgary, Canada; Alberta’s Tomorrow Project, Cancer Control Alberta, Alberta Health Services, Edmonton, AB. Canada; Institute of Diabetes and Clinical Metabolic Research, University Medical Center Schleswig-Holstein, Kiel. Germany; Department of Radiology, University of Calgary, Calgary, Canada; Division of Endocrinology, Diabetes and Clinical Nutrition, Department of Medicine, University Medical Center Schleswig-Holstein, Kiel. Germany; Core Facility Mass Spectrometry, Medical University of Graz, Graz, Austria

**Keywords:** Gut microbiome, hypertension, metabolomics, tryptophan, inflammation

## Abstract

**Background:** Hypertension affects over 1.28 billion adults worldwide, including a significant number of women. Although the gut microbiome is implicated in the onset and progression of hypertension, few studies have examined the relationship in middle-aged women.

**Methods:** Within an established cohort, we investigated the relationship between gut microbiota and its metabolites in normotensive vs. hypertensive middle-aged women (n=108) matched for age (56.6±0.91 years) and body mass index (24.3±0.24 kg/m²). Fecal microbiota analysis was performed using 16S rRNA sequencing and serum metabolites were analyzed using LC-MS/QTOF. Associations between the microbiota, metabolomic alterations and systemic inflammatory cytokines were statistically examined to uncover their interrelationships and potential role in disease progression.

**Results:** Women with hypertension had gut dysbiosis with an increased Firmicutes/Bacteroidetes ratio and higher abundances of inflammatory taxa including *Anaerostipes* and *Collinsella*. Untargeted serum metabolomics demonstrated that hypertensive participants had elevated levels of tryptophan, the pro-inflammatory metabolite kynurenine and lower levels of health-promoting indoles produced by the action of gut microbiota on tryptophan (p<0.05). These findings were confirmed in microbiota analysis showing a reduced abundance of indole-producing species (*Alistipes shahii*, *Bacteroides faecichinchillae, Bacteroides stercoris*)(p<0.05) suggesting a lower microbial activity of tryptophan-indole metabolism. Furthermore, hypertension increased inflammatory markers including an elevated IL12/IL10 ratio, interferon-γ and tumor necrosis factor-α. The IL-12/IL-10 ratio demonstrated a positive correlation with kynurenine levels, emphasizing the involvement of cytokines and gut microbiota in driving systemic inflammation in hypertension.

**Conclusion:** Imbalances in microbiota-regulated tryptophan metabolism contribute to systemic inflammation in hypertensive, middle-aged women, presenting a potentially modifiable target for intervention.

**GRAPHICAL ABSTRACT:** 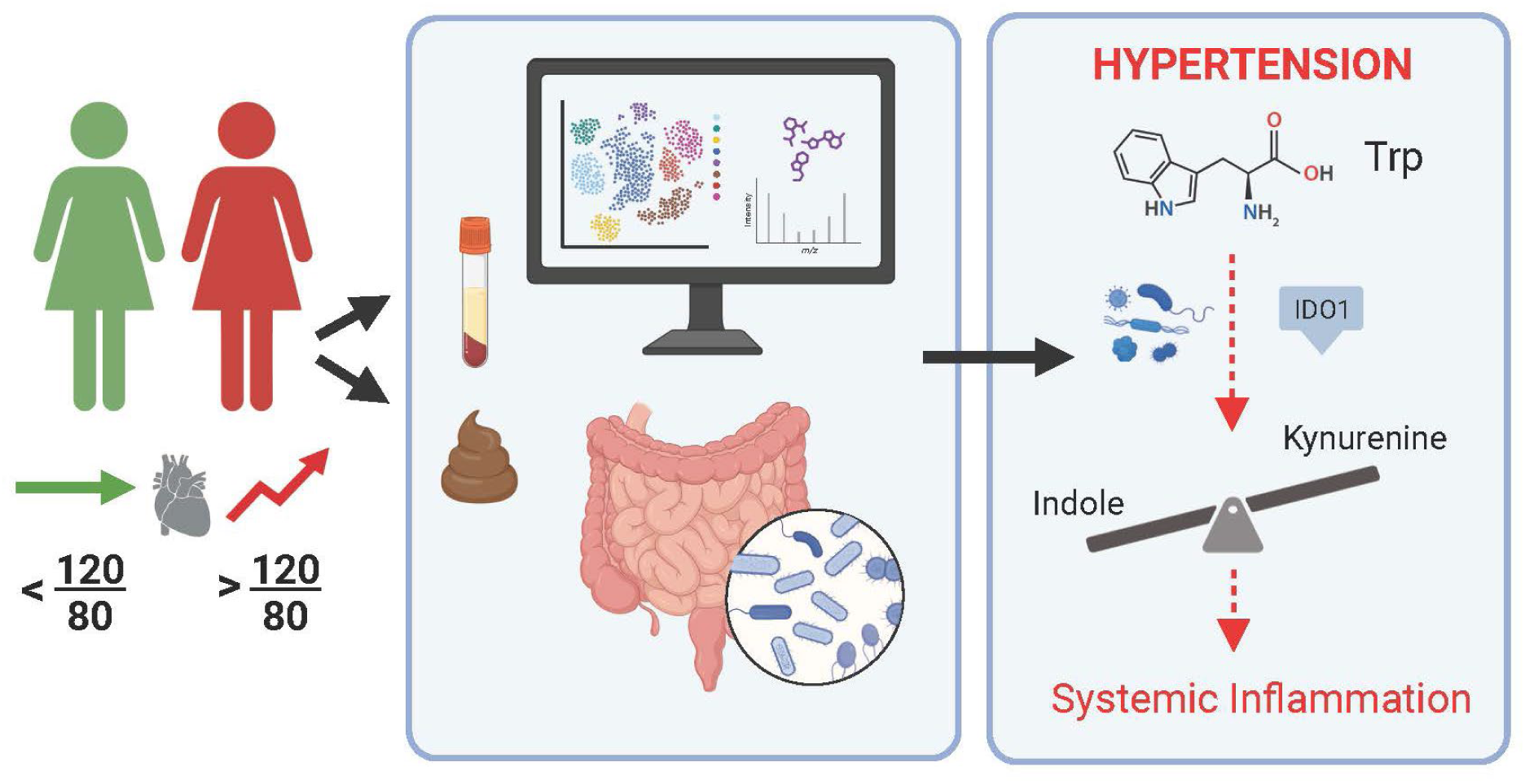

## INTRODUCTION

Hypertension, or elevated blood pressure, is a major contributor to cardiovascular disease and premature death worldwide.^1^ Notably, middle-aged women are at greater risk of developing hypertension due to hormonal changes occurring during and after menopause.^2^ Starting as early as the third decade of life, women experience a steeper increase in blood pressure compared to men, a trend that continues to persist throughout their lives. This rise in blood pressure also comes with a disproportionate risk in women - a 10 mmHg increase in systolic blood pressure in women translates into a 25% higher risk of cardiovascular disease, whereas the comparable risk for men is 15%.^3^ As the most prevalent modifiable risk factor for cardiovascular disease progression, understanding the factors contributing to hypertension in women is crucial.

Beyond pathophysiologic mechanisms, various environmental elements impact the risk, advancement, and severity of hypertension. These include physical activity, diet, socioeconomic status, and the gut microbiota - the trillions of microbes inhabiting our intestinal tract. It is now known that the gut microbiota plays a causal role in blood pressure regulation, as demonstrated by the transfer of hypertensive phenotypes through fecal transplant in several studies.^4–6^ Microbial disruptions with hypertension have been documented in numerous cohort studies^7–9^ to date including the Twins UK study where women had reductions in both alpha and beta diversity, as well as distinctive microbial signatures compared with their normotensive counterparts.^10^ One mechanism by which the gut microbiota may regulate blood pressure is through its bioactive metabolites.^11,12^ Of particular interest to the present study is a class of metabolites produced by the action of gut microbes on dietary tryptophan, an essential amino acid obtained from protein-rich foods and metabolized in the gut by host enzymes or by the action of proteolytic bacteria.^13–16^

Upon reaching the colon, a fraction of tryptophan (1%) is utilized for protein synthesis, while the remainder is catabolized into kynurenine (∼95%), indoles (2%), or serotonin (1-2%) via their respective pathways.^11^ Among these, kynurenine and its downstream metabolites are diagnostically and prognostically implicated in cardiometabolic disease.^17^ Increased levels of kynurenine pathway metabolites are associated with adverse hemodynamics^18^ as well as hypertension prediction and mortality.^19^ In contrast, indoles produced from the actions of gut microbiota on tryptophan, have been extensively reported in maintaining peripheral health by fine-tuning intestinal physiology, anti-inflammatory, and antioxidant potential.^16,20^ Although the role of indoles in hypertension is controversial, reduced indole levels have been reported in metabolic disease states^21–23^ while indole propionic acid has been demonstrated to modulate cardiomyocyte metabolism and to reduce systolic blood pressure and inflammation in a model of salt-sensitive hypertension.^24^

This study aims to elucidate the relationship between gut microbiota, tryptophan metabolites, and blood pressure in age and BMI-matched women (40-65 years, 18.5-29.9 kg/m^2^) with hypertension. Matching is essential as excess body weight often accompanies hypertension in this population. Furthermore, given the direct link between gut microbiota and systemic inflammation^25^, the association between inflammatory cytokines and gut-regulated tryptophan metabolites in normal and hypertensive women was investigated. It was hypothesized that alterations in gut-derived metabolites, particularly tryptophan metabolites might be associated with dysbiosis and systemic inflammation in hypertensive participants compared to their normotensive counterparts.

## METHODS

### Study Population, Sample Collection and Anthropometric Measures

The Conjoint Health Research Ethics Board at the University of Calgary (REB17-1973) granted approval for the study. A total of 175 women between the ages of 38-65 years were re-recruited from the Alberta’s Tomorrow Project cohort (Calgary, AB, Canada)^26,27^ based on their age, BMI, medical history, and health status. Out of a total of 77 eligible women: n = 19 were diagnosed with other cardiovascular conditions (non-HT), and n= 4 were smokers who were excluded. Pregnant women, cancer patients and women who used antibiotics in the past 3 months were also excluded. Eligible women were clinically diagnosed with hypertension by their physician within the year prior to recruitment (n = 54). Hypertensive women were age- and BMI-matched to normotensive women (n = 54) given the direct involvement of these factors in hypertension.^28,29^ Details on eligibility criteria and participant selection is illustrated in **Supplementary Figure 1**.

Dietary and lifestyle surveys were emailed to eligible participants once written informed consent was obtained.^26^ Detailed information on the questionnaires can be found in the following link - https://myatp.ca/resources/previous-surveys. Subsequently, a fecal collection kit (Protocult 120, Abilities Build Products, USA) was mailed with instructions on how to collect samples at home. Participants stored the collected stool sample at -20°C and brought the sample to the study site within 48 hours. On the day of the appointment, participants arrived at the research facility following an overnight fast, and a trained phlebotomist obtained blood via an antecubital puncture and serum isolated. Collected samples were then stored at -80°C until further processing. During the visit, the measurements described below were also obtained by trained staff at the study site: standing/sitting height (cm), weight (kg), systolic/diastolic blood pressure (mmHg), heart rate (bpm), waist-hip circumference (cm), and hand grip strength (kg). Details of these procedures have been previously published.^26,27^

### Inflammatory Biomarkers

To examine the inflammatory status of the study participants, traditional cardiovascular disease biomarkers and cytokines were analyzed using the *Human Cardiovascular Disease Panel 3 9-Plex Discovery Assay® Array, HDCVD15,* and *Human Cytokine Proinflammatory Focused 15-Plex Discovery Assay® Array, HDF15* (MILLIPEX Eve Technologies Corporation, Canada), respectively. The cardiovascular disease panel consisted of α2-Macroglobulin, α1-acid glycoprotein (AGP), C-reactive protein (CRP), Fetuin A, Fibrinogen, Haptoglobin, L-Selectin, platelet factor (PF4), and Serum Amyloid P component (SAP). The cytokine panel measured GM-CSF, interferon γ (IFN-γ), interleukin-(IL-) 1β, IL-1RA, IL-2, IL-4, IL-5, IL-6, IL-8, IL-10, IL-12(p40), IL-12(p70), IL-13, monocyte chemoattractant protein 1 (MCP-1), and tumor necrosis factor α (TNF-α).

### Fecal DNA Extraction, Amplification, and Sequencing

For gut microbial diversity and composition, total genomic DNA was extracted from fecal samples using the QIAamp Fast DNA stool mini kit (Qiagen, Germany) as described previously^30^. Briefly, fecal samples (∼200 mg each) were mixed with InhibitEX buffer, homogenized using a SpeedMill PLUS (Analytic Jena, Germany), heated at 95°C for 5 minutes, and centrifuged for 60 seconds. The resulting supernatant was processed further for automated DNA isolation, following the manufacturer’s protocol. Quality of the DNA was checked using Nanodrop 2000 Spectrophotometer (Thermo Fisher Scientific, USA). The purified DNA pellets were stored at - 20°C until further use. Samples were sequenced following amplification of the V3−V4 hypervariable region of the 16S rRNA gene on a MiSeq platform (Illumina, USA) as previously described.^31^

Preprocessing of the 16S rRNA data was conducted in R (v.4.1.3) using *phyloseq* package (v.1.34.0) followed by quality control, threshold filtering, and rarefaction steps.^30^ This rarefied dataset was used for the analysis unless otherwise specified. Alpha and beta diversity were calculated using the Shannon Diversity Index^31^ and Bray-Curtis dissimilarity metrics^32^, respectively. Relative abundances were center log-ratio (CLR) transformed following zero-replacement using *zCompositions* (v.1.3.4) and *CoDaSeq* (v.0.99.6) to control for composition before statistically assessing differential abundance. Demultiplexed sequences generated for this study are deposited on the NCBI Sequence Read Archive (SRA PRJNA922681).

### Serum Metabolomics

To determine a broad range of aqueous as well as non-aqueous serum metabolites, untargeted metabolomics, and lipidomics were performed separately. Samples for protein precipitation and metabolite extractions were prepared using LC/MS grade methanol. Briefly, 50 µl of the serum sample was mixed with 200 µl of ice-cold 100% methanol (5 µM) containing internal standard, sonicated, and centrifuged at 14,000 rpm (10 min). The supernatant was collected and placed in the Speed-Vac system at 45°C until completely dry. Extracts were then reconstituted using 50% methanol, centrifuged, and filtered twice using 0.2 µM filter to remove any impurities. Samples were analyzed using Agilent 6550 iFunnel Q-TOF LC/MS system (Agilent, Santa Clara, CA, USA). Metabolites were separated using Acquity UPLC HSS T3 C18 column (2.1×100mm, 1.8um size; Waters Corporation, USA) in gradient mode (Mobile Phase → H_2_O, Mobile Phase → 100% Acetonitrile). Data processing and machine parameters are described in detail elsewhere.^33^ To check the reproducibility, quality control samples were run after every 12 samples. Raw data were converted using ProteoWizard 4.0 software to mzXML format and uploaded to XCMS online.^34^ Over 20,000 different serum metabolites were detected. Data were normalized using median-fold change and auto-scaling features in MetaboAnalyst 5.0 (https://www.metaboanalyst.ca/) for the extracted peak abundance^35^. Metabolites were then identified using Human Metabolome Database, HMDB (http://www.hmdb.ca/)^36^, PubChem (https://pubchem.ncbi.nlm.nih.gov) ^37^, and METLIN databases (http://metlin.scripps.edu).^38^

### Serum Lipidomics

For quantification of non-aqueous metabolites, serum samples were extracted using a modified Matyash method^39^, and obtained lipid extracts were further prepared for positive and negative ion electrospray ionization separately as previously described.^40^ Samples were stored at -80 °C until further analysis. Next, mass spectrometry was performed using a Q-Exactive Focus hybrid quadrupole-orbitrap mass spectrometer coupled to a Vanquish UHPLC system (both ThermoFisher, USA). Detailed instrument parameters and a data processing and analysis workflow have been previously published.^40^ Evaluation of mass features in lipidomics analysis was conducted using DataAnalysis 5.0 and MetaboScape 4.0.1 both from Bruker (Bremen, Germany). Downstream analysis of the identified lipid species was performed using MetaboAnalyst 5.0.^35^

### Statistical Analysis

Categorical data are presented as mean ± SE unless otherwise indicated. Differences between normotensive and hypertensive participant characteristics, cytokine levels, and cardiovascular disease markers were analyzed using unpaired t-tests with Welch’s correction applied where appropriate. Outliers were identified and removed using the ROUT method in GraphPad Prism v10 (Boston, USA).^41^ Statistical significance was set at p ≤ 0.05, and where applicable multiple testing correction was performed using the false discovery rate (FDR), with an adjusted threshold of q < 0.25.

For gut microbiota data, alpha diversity differences between groups were assessed using the Kruskal-Wallis test and beta diversity differences were evaluated using Permutational Multivariate Analysis of Variance (PERMANOVA) with 999 permutations, as implemented in the ‘adonis2’ function from the *vegan* package (v2.5-7) in R (v4.1.0). Redundancy analysis was performed using *vegan* (v. 2.5-4) to extract and summarize the variation in the entire gut microbiota data^42^ by accounting for potential confounders - BMI, diet, age, and physical activity. Metabolomics data were analyzed using partial least squares-discriminant analysis (PLS-DA) to assess group separation, followed by variable importance in projection (VIP) scores to rank metabolites by their contribution to discrimination. Metabolites with both a VIP score >2 and *p* < 0.05 were considered significantly different. Pathway enrichment and metabolic pathway analyses were conducted using MetaboAnalyst v5.0^35^, with statistical significance assessed via Fisher’s exact test. For lipidomics data, PLS-DA was used to identify discriminant lipid species across the study groups. Significant lipid species were further analyzed using a volcano plot to visualize fold changes and p-values. Lipid species were considered significant if they met a threshold of p < 0.05 and FDR-adjusted q < 0.25.

Multivariate associations were performed using a machine-learning approach, *Data Integration Analysis for Biomarker discovery using Latent Components* (DIABLO, *mixOmics* package in R, v.6.19.4) to understand the relationship between physical characteristics (BMI, systolic/diastolic BP, heart rate, age, and diet), altered inflammatory markers (AGP, CRP, SAP, INF-γ, TNF-α, PF4, IL5, IL10, and fetuin A), gut microbiota, and tryptophan metabolites. Data for multivariate analysis was internally cross-validated and a correlation cut-off of 0.7 was set to identify the most robust associations.^43^

## RESULTS

### Participant Characteristics

A total of 108 middle-aged women matched for age and BMI were included in the study. Participant characteristics are shown in **Table 1**. There were no differences in anthropometric measures between participant groups including age (years), height (m), weight (kg), BMI (kg/m^2^), or hand-grip strength (kg). Compared to normotensive participants, those clinically diagnosed with hypertension had higher systolic and diastolic blood pressure (*p* < 0.0001) (**Figure 1A**) with no significant differences in resting heart rate (*p* = 0.40) (**Table 1**). It should be noted that 76% of hypertensive participants were on prescribed medication(s) (**Table 1**) and yet 33.3% had elevated blood pressure values. 22.2% were classified as Stage 1 hypertension, and 24.1% as Stage 2 hypertension according to the classification by the American Heart Association (www.heart.org/bplevels). When blood pressure medication was accounted for by adding 15 mmHg and 10 mmHg to the systolic and diastolic blood pressure^44^, even greater differences between groups emerged (**Figure 1A**).

**Table 1.** Participant Characteristics. Data is presented as mean ± SEM. Values in bold indicate significant differences between groups (*p*<0.05). Abbreviations: ACE inhibitors; angiotensin-converting enzyme inhibitors; ARBs, angiotensin II receptor blockers; BMI; body mass index, bpm; beats per minute; CCBs, calcium channel blockers; HR; heart rate.

|  | Normotensive<br>(n= 54) | Hypertensive<br>(n=54) | P-value |
| --- | --- | --- | --- |
| Age (years) | 55.69 ± 0.91 | 57.51 ± 0.91 | 0.112 |
| Height (cm) | 87.68 ± 0.37 | 87.52 ± 0.47 | 0.790 |
| Weight (kg) | 65.29 ± 0.66 | 66.16 ± 1.31 | 0.554 |
| BMI (kg/m<sup>2</sup>) | 23.99 ± 0.11 | 24.50 ± 0.17 | 0.201 |
| Waist: Hip ratio | 0.86 ± 0.01 | 0.87 ± 0.01 | 0.860 |
| Resting heart rate (bpm) | 63.63 ± 0.96 | 64.96 ± 1.05 | 0.402 |
| Systolic blood pressure (mmHg) | **114.15 ± 1.72** | **130.19 ± 1.77** | **<0.0001** |
| Diastolic blood pressure (mmHg) | **70.24 ± 1.09** | **79.02 ± 1.18** | **<0.0001** |
| Diet score (mMDS) | **4.44 ± 1.71** | **3.73 ± 1.80** | **0.050** |
| Physical activity score (CDHQII) | 267.72 ± 25.16 | 307.04 ± 28.21 | 0.345 |
| Hypertensive Medication (n) | 0/54 | 41/54 |  |
| ACE inhibitors | - | 13 |  |
| ARBs | - | 11 |  |
| Beta blockers | - | 5 |  |
| CCBs | - | 7 |  |
| Diuretics | - | 14 |  |
| Combined Medications | - | 9 |  |

**Figure 1.**
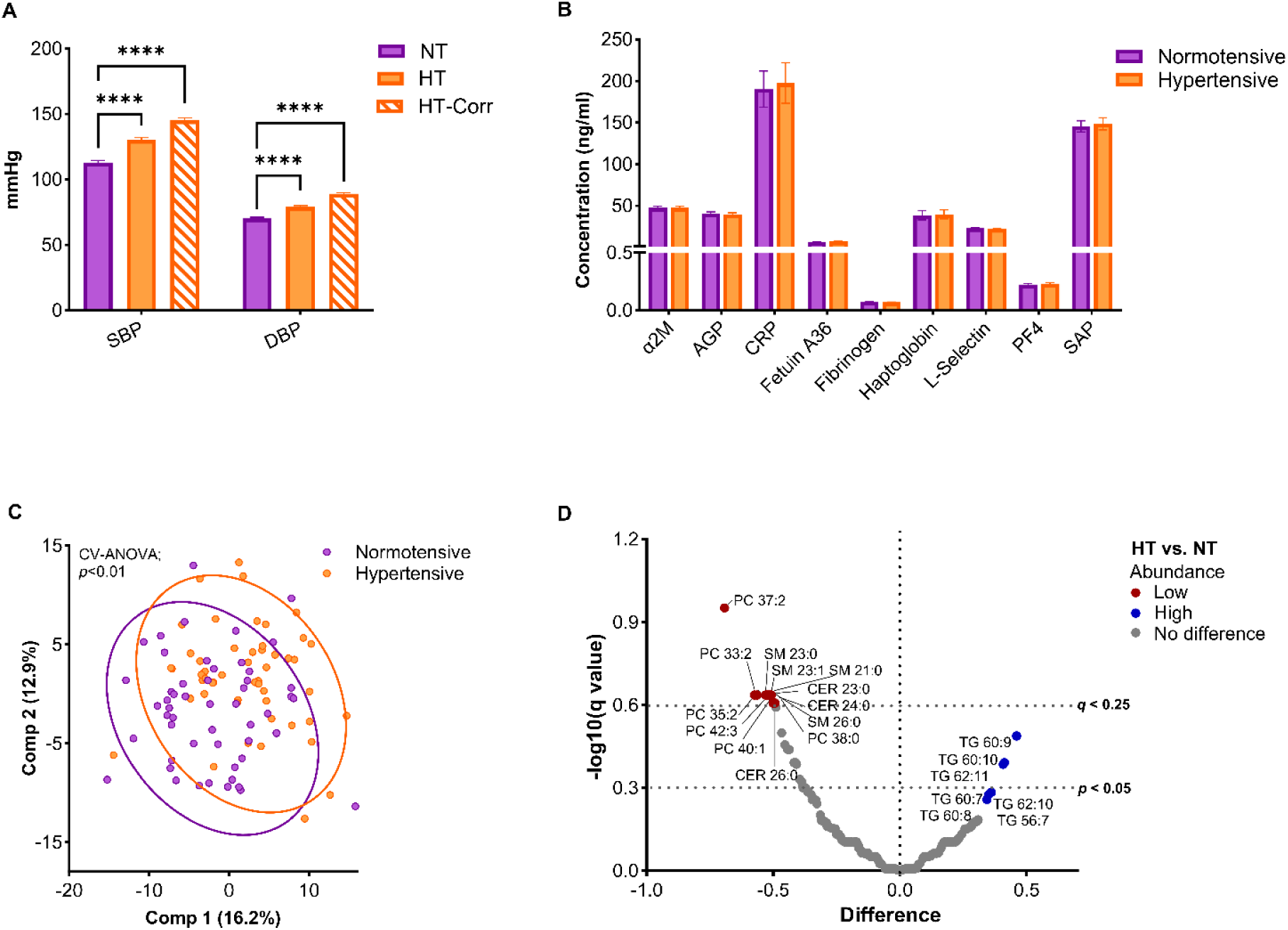
Phenotypic characteristics associated with hypertension. A) Systolic and diastolic blood pressure in study participants. Third bar (hatched) accounts for the theoretical influence of blood pressure medication in hypertensive individuals. 15 mmHg and 10 mmHg were added to the systolic and diastolic blood pressure respectively. B) Cardiovascular disease biomarkers (ng/ml), C) PLS-DA of the serum lipids, and D) Volcano plot showing upregulated vs. downregulated lipid species between HC vs. HT participants. All data are presented as mean ± SEM. *p* < 0.0001 is indicated by ****. Abbreviations: α2M, α2-macroglobulin; AGP, alpha glycoprotein; CE, cholesteryl ester; CER, ceramide; CRP, C-reactive protein; CV-ANOVA, cross-validation analysis of variance; DBP, diastolic blood pressure; HT, hypertensive; NT, normotensive; PC, phosphotidylcholine; PF4, platelet factor 4; SAP, serum amyloid P component; SBP, systolic blood pressure; SM, sphingomyelin; TG, triacylglycerol.

There were no differences in cardiovascular biomarkers between the normotensive vs. hypertensive participants (**Figure 1B**) suggesting early-stage cardiometabolic disease in most the participants. However, participants with hypertension displayed distinct serum lipid profiles (**Figure 1C**) with a lower concentration of phosphatidylcholine (PC - 37:2, 33:2), sphingomyelin (SM - 21:0, 23:0, 23:1), and cholesteryl esters (CE - 23:0, 24:0) and higher concentration of triacylglycerols (TG - 62:11, 60:10, 60:9) in hypertensive compared to normotensive women (**Figure 1D**). Participants with hypertension also had suboptimal dietary habits as calculated by a modified Mediterranean diet score^45^ (*p* < 0.05) with a significant lower daily intake of dietary fiber (*p* = 0.02) (**Supplementary Table 1**).

### Imbalance in the gut microbial composition in hypertension

To elucidate associations of gut microbial composition with hypertension, a comprehensive evaluation of microbial diversity and abundance across phylum, genus, and species levels was conducted. There were no differences in alpha or beta diversity metrics between the study groups (*p*>0.05)(**Figures 2A and B**). Redundancy analysis revealed that uncorrected blood pressure contributed most to the variability in microbial composition (explained variance, r= 0.11 (13.6%), *p*=0.008), followed by BMI (r=0.094 (8.9%), *p*=0.001) and diet (r=0.089 (8%), *p*=0.06) variables (**Figure 2C**). Differential abundance analysis at the phylum level showed a higher abundance of Proteobacteria (*p* = 0.03), and Firmicutes: Bacteroidetes (F/B) ratio (*p* = 0.02) with a decreased abundance of Actinobacteria (*p* = 0.01) in women with elevated BP compared to their normotensive counterparts (**Figure 2D**). At the genus level, hypertensive participants showed an increase in *Anaerostipes* (*p* = 0.01), *Blautia* (*p* = 0.01), and *Collinsella* (*p* = 0.02) genera and a decrease in abundance of *Parasutterella* (*p* = 0.04), *Phascolarctobacterium* (*p* = 0.008), and *Subdoligranulum* (*p* < 0.005) genera (**Figures 2E**).

**Figure 2.**
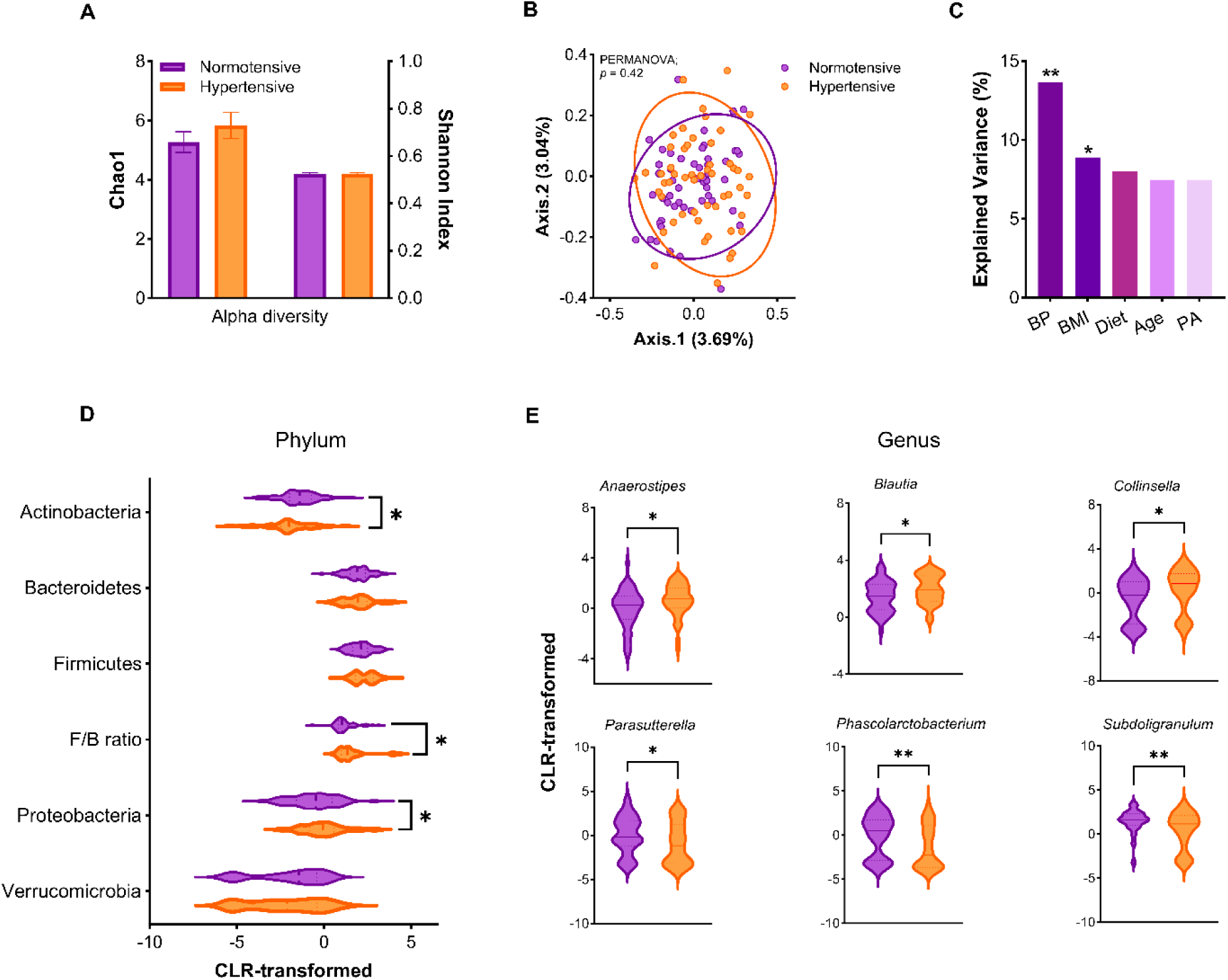
Fecal microbial profile of normotensive vs. hypertensive participants. A) Alpha diversity; B) Beta diversity; C) Redundancy analysis plot; CLR-transformed abundance of D) Phyla; and E) Genus. Unpaired t-test was performed, and statistical difference of *p* < 0.01 and *p* < 0.05 is indicated by ** and *, respectively. Abbreviations: BP, blood pressure; BMI, body mass index, CLR, center-log ratio; F/B ratio, Firmicutes: Bacteroidetes ratio; PA, physical activity; PERMANOVA, permutational analysis of variance.

### Gut-mediated tryptophan metabolism is altered during hypertension

Supervised clustering analysis of serum metabolites by PLS-DA showed a distinct separation between the study groups (CV-ANOVA, *p* < 0.0001) (**Figure 3A**). The top 15 metabolites contributing to the separation between groups are shown in **Figure 3B**. Pathway analysis of all identified metabolites between study groups showed the top hits to include tyrosine metabolism (impact = 0.78, p=0.06), tryptophan metabolism (impact = 0.96, p=0.1), and linoleic acid metabolism (impact = 6.6, p=0.1) (**Figure 3C**).

**Figure 3.**
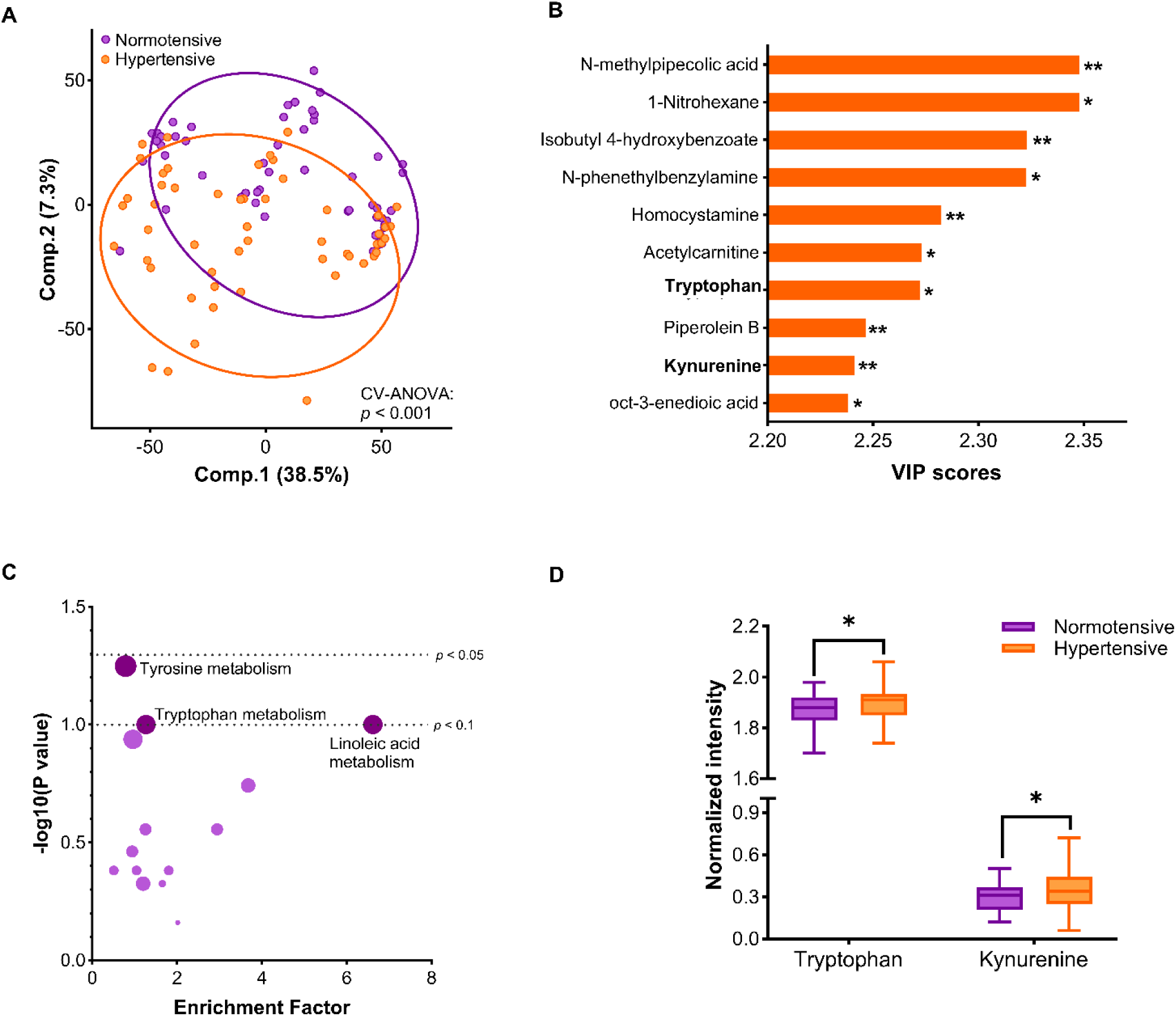
Serum metabolomic profile of normotensive vs. hypertensive participants. A) PLS-DA plot; B); Pathway enrichment analysis; C) VIP score plot of most significantly altered metabolites (microbiota-associated metabolites are highlighted in bold); serum concentration of D) Tryptophan and Kynurenine. Statistical difference of *p* < 0.01 and *p* < 0.05 is indicated by ** and * respectively. All data are presented as mean ± SEM. Unpaired t-test was performed, and *p* ≤ 0.05 was considered significant. Abbreviations: PLS-DA, Partial Least Squares Discriminant Analysis; VIP, variable importance in projection.

Among the altered metabolites in normotensive vs. hypertensive participants, tryptophan and kynurenine were of particular interest because of their association with gut microbiota.^46,47^ Univariate statistical analysis of tryptophan and its metabolites revealed a significant increase in tryptophan and kynurenine (**Figure 3D**) in hypertensive versus normotensive women. However, there were no changes in serotonin or other tryptophan metabolites including kynurenic acid, indole propionic acid, 5-hydroxyindole, tryptamine, melatonin, or the *Kyn:Trp* ratio (**Supplementary Figure 2**).

### Lower abundance of indole-producing bacteria in hypertension

Given that indole, a tryptophan metabolite is primarily produced by gut microbiota^20^, we then investigated indole concentration and the differential bacterial species that encode indole-producing enzymes, tryptophanase (*Trp*A) or indole-3-glycerol phosphate synthase (*Trp*C). Interestingly, women with hypertension had lower levels of circulating serum indole (**Figure 4A**) and a decreased abundance of *Alistipes shahii* (*p =* 0.03), *B. faecichinchillae*, and *B. stercoris* (*p* = 0.02) compared to their normotensive counterparts (**Figure 4B**). Overall, metabolomics results show that tryptophan metabolism is altered in hypertension with an increase in host-regulated kynurenine and decreased production of gut microbiota-derived indole.

**Figure 4.**
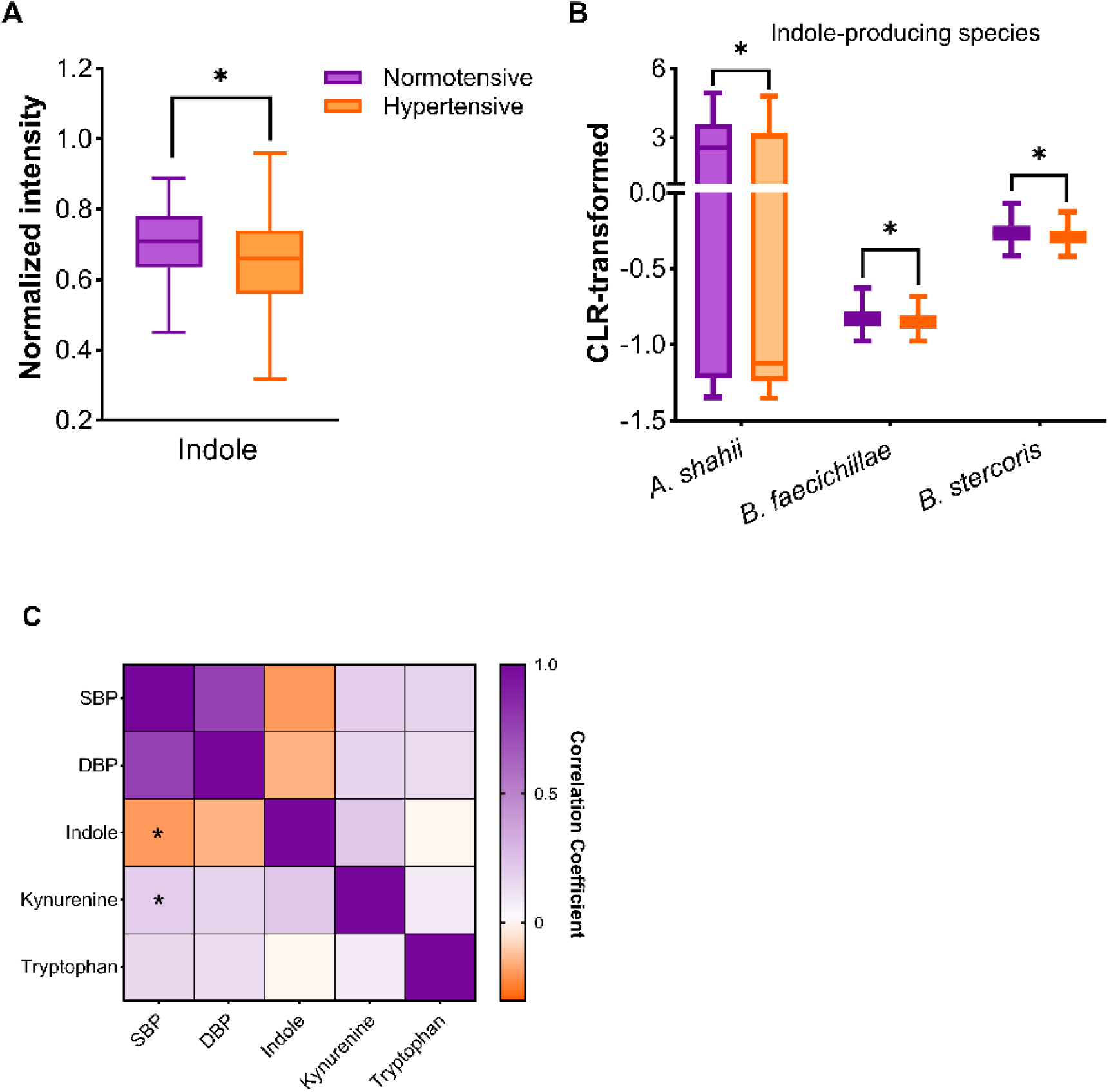
Association of indole-producing bacterial species with hypertension. A) Serum concentration of Indole; B) Relative abundance of indole-producing species; and, C) Spearman correlation between tryptophan metabolites and blood pressure. Statistical difference of *p* < 0.05 is indicated by *. Abbreviations: *Alistipes shahii*, *Bacteroides faecichinchillae*, *Bacteroides stercoris*.

To explore the potential relationship between tryptophan metabolites with blood pressure, a correlation matrix was further constructed. As shown in **Figure 4C**, the serum indole was negatively correlated with systolic blood pressure (*p* < 0.05), while kynurenine was positively correlated with the same, indicating the potential involvement of tryptophan metabolites on blood pressure regulation.

### Cytokine profiles and their associations with metabolites

To understand immune response and potential immunometabolic implications in hypertension, blood cytokines were determined, as well as their correlations with metabolites. Hypertensive participants had higher levels of inflammatory cytokines such as INF-γ, TNF-α, and IL12/IL10 ratio compared to normotensive participants (**Figure 5A**). Concentrations of other cytokines and chemokines in blood, including IL-4, IL-5, IL8, IL13, and MCP-1, were similar between normotensive and hypertensive participants (**Supplementary Table 2**). Correlation analysis showed a significant positive correlation between the concentration of serum kynurenine and IL12/IL10 *(p* = 0.02, Spearman’s rho = 0.05) (**Figure 5B**). Additionally, multivariate canonical correlation using DIABLO revealed other proinflammatory cytokines (IFN-γ, IL12/IL10, TNF-α), and kynurenine:tryptophan (*Kyn:Trp*) ratio showed positive associations with systolic as well as diastolic blood pressure (r ≥ 0.7), indicating the potential role of these metabolites in regulating immune response during hypertension (**Supplementary Figure 3**). A schematic diagram summarizing the results is shown in **Figure 5C**.

**Figure 5.**
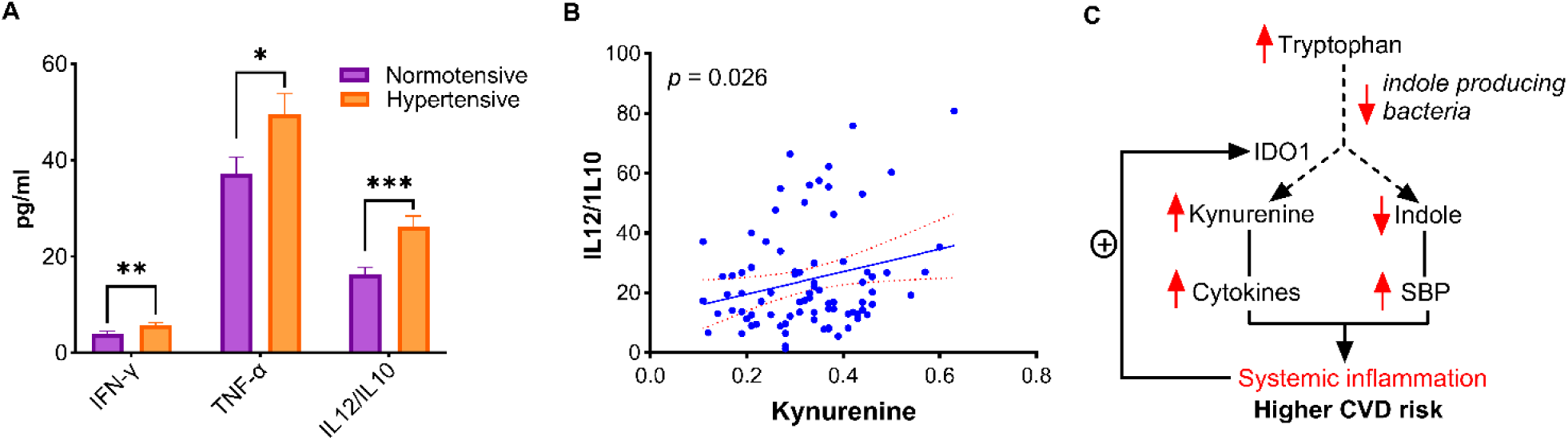
Inflammatory profile of normotensive vs. hypertensive participants. A) Blood cytokines; B) Simple linear regression between IL12/IL10 ratio and kynurenine; and C) Schematic diagram showing tryptophan metabolism during hypertension (red arrows represent increase or decrease in metabolite concentrations in normotensive vs. to hypertensive participants). Statistical difference of *p* < 0.001, *p* < 0.01, and *p* < 0.05 is indicated by ***, **, and * respectively. Abbreviations: IDO1, indolamine-2,3 dioxygenase 1; IFN-γ, interferon-gamma; IL, interleukin; TNF-α, tumor necrosis factor-alpha.

## DISCUSSION

Emerging research implicates gut microbiota as a potential modulator of cardiovascular health and disease. Despite considerable research in the field, the etiology of hypertension is poorly understood in women of menopausal age and there is limited knowledge about the role of the gut microbiota and its metabolites. To address this gap, fecal microbiome and serum metabolomics analyses were performed in a cohort of age and BMI-matched middle-aged women along with cardiovascular and immunological profiling. Compared to the normotensive women, those with hypertension displayed gut dysbiosis with blood pressure being the largest contributor to gut microbiota variability. These alterations, characterized by disrupted gut-mediated tryptophan metabolism and significantly elevated levels of circulating inflammatory biomarkers, strongly implicate the gut microbiota as a key contributor to the development and perpetuation of hypertension in middle-aged women.

Dysregulation of lipid metabolism is a key feature of hypertension.^48,49^ Although women in this study were age and BMI matched to reduce the risk of these confounders, there were notable differences in serum lipids between women with and without hypertension. Hypertension was associated with lower PC, SM, and CE lipid species and higher TG, phenotypes that would be expected to contribute to the progression of cardiometabolic disease. Of particular interest, PC is the precursor to gut microbiota-derived trimethylamine N-oxide (TMAO), a metabolite well known to predict cardiovascular disease risk^50^. Increased PC conversion resulting in decreased circulating PC levels including PC34:2 and PC36:5 have been previously reported in pulmonary hypertension.^51^ Although different PC species (PC37:2 and PC33:2) were decreased in the present study, these findings underscore the potentially crucial role of PC in the pathogenesis of hypertension.

Hypertension is a complex condition with a multifactorial etiology influenced by a combination of genetic, dietary, environmental and behavioral factors. Of these factors, diet and microbiota play a crucial role in shaping metabolic phenotypes linked to hypertension and are even suggested to have a greater impact on hypertension susceptibility than genetic factors.^52^ Comprehensive microbiota profiling identified a notable increase in Firmicute/Bacteroidetes ratio and Proteobacteria in the participants with hypertension. An increase in Firmicute/Bacteroidetes ratio has also been found in individuals with hypertension^53^ and male rats with hypertension induced by a high-salt diet.^54^ Similarly, an increase in Proteobacteria was also reported in a hypertension cohort including both men and women.^55^ Notably, a significant blooming of Proteobacteria was further observed in hypertension patients with cognitive impairment.^56^ While the exact mechanisms by which these alterations are associated with hypertension remain unclear, these phylum-level changes suggest a shift in microbial structure that may be indicative of hypertension status.

At the genus level, microbial alterations in the present cohort revealed a unique gut microbiota composition in hypertensive women characterized by an increase in *Anaerostipes* and *Blautia*, and a decrease in *Phascolarctobacterium* and *Parasutterella*. Of these, the *Blautia* finding is counter to what has been previously observed. Notably, *Blautia* species have been shown to be depleted in hypertension compared to matched controls.^55^ However, there are also clear sex specific differences in gut microbiota dysregulation associated with blood pressure. Of note, Virwani *et al.* report a positive association between *Blautia hansenii* and SCFA in women with hypertension, but no relationship in men.^57^ Combined, these findings indicate that there may be sex specific differences in the gut microbiota that contribute to the pathology of hypertension.

Given the observed alterations of fecal microbiota, the question arises whether these microbial shifts are reflected in metabolic functions. Non-targeted high-resolution serum metabolomics identified alterations in several metabolites associated with hypertension, including tryptophan metabolites (tryptophan, kynurenine, and indole), LysoPC (16:0), and acetylcarnitine. Tryptophan metabolism is particularly intriguing due to its known role in cardiovascular disease.^58^ In this study, elevated levels of tryptophan and kynurenine were found in hypertensive participants. Kynurenine is produced in host cells via indoleamine 2,3 dioxygenase (IDO1), which is increased in a myriad of inflammatory states including obesity^23^, heart failure^59^ and cardiovascular disease.^60^ Intravenous infusion of kynurenine raises mean arterial pressure and heart rate in rodents^61^, further substantiating its causative role in the pathogenesis of hypertension. Of interest to the present study, gut microbial composition has been implicated in enhancing host tryptophan metabolism, increasing the conversion of tryptophan to kynurenine.^62^

In contrast, circulating indole was reduced in hypertensive women in this study. This metabolite is primarily produced by the action of gut microbes on tryptophan.^63^ The metabolite plays important roles in microbe quorum sensing, intestinal homeostasis and in the modulation of immune homeostasis via the aryl hydrocarbon receptor axis.^64^ The present study found a decrease in microbesinvolved in indole metabolism with hypertension, including *Alistipes shahii* and *Bacteroides stercoris* which encode tryptophanase, a rate-limiting factor in the biosynthesis of indole. Studies in rat models have shown that indole acts as a hemodynamic modulator and can reduce arterial blood pressure when administered intracerebroventricularly.^65^ Interestingly, a negative correlation was observed between indole and systolic blood pressure in this study, indicating a regulatory role of indole that warrants further investigation.

Accompanying the shift in tryptophan metabolism, women with hypertension also exhibit an increased production of proinflammatory cytokines, including IFN-γ and TNF-α. The elevated ratio of IL12/IL10 further indicates systemic low-grade inflammation. These inflammatory alterations have been linked to impaired insulin sensitivity, vascular calcification, macrophage migration as well as, lipid accumulation in blood vessels, liver, and adipose tissue.^15,66–70^ IFN-γ is known as a stimulator of IDO1 expression,^71^ which drives the metabolism of tryptophan towards kynurenine. IL12 could also promote IDO1 expression while IL10 exerts inhibitory effects.^72^ In the present study, the positive correlation between IL12/IL10 and kynurenine underscores the role of inflammation in modulating tryptophan-kynurenine metabolism in the context of hypertension.

This study provides valuable insights into the connection between gut microbiota, metabolites, and hypertension in middle-aged women, though it is not without certain limitations. The lack of racial and ethnic diversity in the cohort restricted to a specific demographic of age- and BMI-matched women may limit the generalizability of the findings to broader populations.^73^ Next, the use of 16S rRNA sequencing for microbiome analysis provides taxonomic resolution but lacks the depth of functional profiling achievable through shotgun metagenomics.^74^ Finally, while associations were identified, the observational design of this study precludes causal inferences, emphasizing the need for longitudinal studies or controlled interventions to validate these findings.

This study highlights the role of altered microbiota-dependent tryptophan metabolism in the pathogenesis of hypertension in middle-aged women. Elevated levels of proinflammatory kynurenine, alongside a reduction in anti-inflammatory indole - primarily driven by gut microbiota dysbiosis - contribute to a state of low-grade systemic inflammation. This inflammatory environment not only worsens hypertension but also substantially increases cardiovascular disease risk. In summary, imbalances in key metabolites, induced by dysbiosis, reveal a complex interplay between microbial metabolism, immune regulation, and vascular health, emphasizing the gut microbiota as a mediator in the development of hypertension and associated cardiometabolic risk.

### Perspectives

This study uncovers novel insights into the gut microbiota’s role in hypertension among middle-aged women, an often overlooked and underdiagnosed group. The findings emphasize the gut microbiome’s influence on tryptophan metabolism, specifically the disruption of the kynurenine and indole pathways, which may drive inflammation and heighten cardiovascular risk in this demographic. The observed links between microbial metabolites and pro-inflammatory cytokines underscore the potential of targeting the gut microbiome as a therapeutic strategy for hypertension. Future research should prioritize clinical trials exploring microbiota modulation such as dietary interventions or targeted probiotics to assess whether restoring microbial balance can effectively reduce the risk of hypertension and related cardiometabolic conditions.

## Novelty and Relevance

### What Is New?

- Compared to age- and BMI-matched normotensive middle-aged women, individuals with mild hypertension exhibit gut microbiota dysbiosis, marked by an increased Firmicutes/Bacteroidetes ratio and Proteobacteria.
- In hypertension, altered gut microbiota disrupts tryptophan metabolism, increasing kynurenine while decreasing indole production.
- Microbiota-induced inflammatory pathways contribute to hypertension, with kynurenine positively correlated with cytokines and indole negatively associated with blood pressure.

### What Is Relevant?

This study unveils novel insights into the relationship between the gut microbiota and hypertension, shedding light on its impact in middle-aged women - one of the highest-risk groups for cardiovascular disease.

### Clinical and Pathological Implications

Microbiota-targeted therapies have the potential to mitigate inflammatory and metabolic dysregulation in the development and progression of hypertension.

## Data Availability

The 16S rRNA gene-sequences data supporting the conclusions of this article are deposited at the National Center for Biotechnology Information (BioProject SRA PRJNA922681).

## Acknowledgement

We would like to sincerely thank the Alberta Tomorrow Project research staff at Richmond Diagnostics Center, Calgary, AB for all their help in participants’ recruitment and follow-ups, sample collection at the facility, data transfer, and coordinating with the research team at University of Calgary. Alberta’s Tomorrow Project is only possible because of the commitment of its research participants, its staff, and its funders: Alberta Health, Alberta Cancer Foundation, Canadian Partnership Against Cancer, Health Canada, and Alberta Health Services. The views expressed herein represent the views of the author(s) and not of Alberta’s Tomorrow Project or any of its funders.”

## SUPPLEMENTARY DATA

**Supplementary Figure 1.**
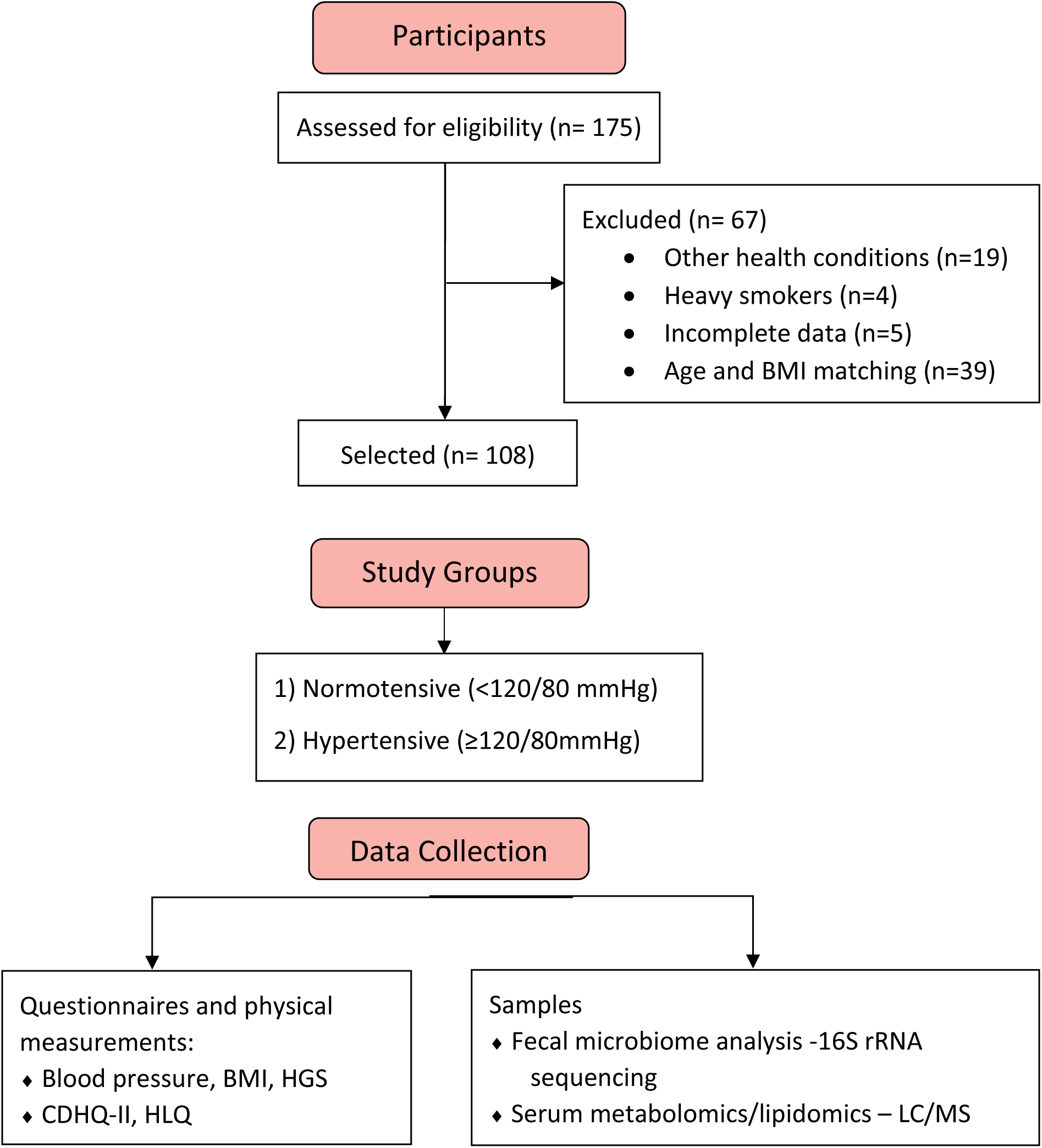
Flowchart of participants selected from Alberta’s Tomorrow Project Cohort and their classification into study groups based on their systolic/diastolic blood pressure and BMI. Abbreviations: BMI, body mass index; CDHQ-II, Canadian Dietary Health Questionnaire; HGS, hand grip strength; HLQ, Health Lifestyle Questionnaire; LC/MS-QToF, liquid chromatography/mass spectrometry-Quadrupole Time of Flight; SBP/DBP, systolic/diastolic blood pressure.

**Supplementary Table 1.** Dietary intake of normotensive vs. hypertensive women. Diet score is based on the Canadian Dietary Health Questionnaire (CDHQ II). A modified Mediterranean diet score diet score was calculated based on the weekly intake of nine food groups adjusted by total energy intake; Vegetables, Fruits, Legumes, Grains, Dairy, Meat, Fish, Alcohol and a fatty acid ratio (calculated as the sum of mono- and poly-unsaturated fatty acid divided by saturated fatty acid intake). The diet scores ranged from 0 (least healthy) to 9 (most healthy). Data represents mean ± standard error. Unpaired t-test was performed and p<0.05 was considered significant.

| Diet | Normotensive | Hypertensive | P-value |
| --- | --- | --- | --- |
| Diet score | 4.44 ± 1.71 | 3.73 ± 1.80 | **0.050\*** |
| Carbohydrate (g) | 156.2 ± 8.1 | 156.2 ± 7.8 | 0.660 |
| Dietary fiber (g) | 18.7 ± 1.1 | 15.8 ± 0.9 | **0.021\*** |
| Protein (g) | 62.8 ± 3.5 | 62.3 ± 3.9 | 0.657 |
| Fat (g) | 64.6 ± 4.0 | 60.2 ± 3.5 | 0.159 |
| Calcium (mg) | 896.6 ± 50.7 | 857.0 ± 54.8 | 0.597 |
| Iron (mg) | 12.1 ± 0.7 | 10.6 ± 0.5 | 0.102 |
| Magnesium (mg) | 357.3 ± 19.3 | 318.0 ± 14.9 | 0.114 |
| Phosphorus (mg) | 1134.5 ± 57.0 | 1144.4 ± 67.6 | 0.910 |
| Potassium (mg) | 2946.1 ± 153.4 | 2826.2 ± 145.1 | 0.573 |
| Sodium (mg) | 2118.4 ± 112.1 | 2113.0 ± 140.2 | 0.976 |
| Zinc (mg) | 9.5 ± 0.4 | 9.2 ± 0.5 | 0.678 |

**Supplementary Table 2.** Serum Cytokines in normotensive vs. hypertensive women. Data represents mean ± standard error. Unpaired t-test was performed and p<0.05 was considered significant. Abbreviations: IL, interleukins; MCP, monocyte chemoattractant protein.

| **Cytokines (ng/ml)** | **Normotensive** | **Hypertensive** | **P-value** |
| --- | --- | --- | --- |
| **IL-4** | 2.4 ± 0.7 | 1.0 ± 0.3 | 0.10 |
| **IL-5** | 7.7 ± 0.8 | 7.0 ± 2.4 | 0.35 |
| **IL-6** | 2.7 ± 0.7 | 1.7 ± 0.3 | 0.31 |
| **IL-8** | 11.8 ± 1.1 | 11.4 ± 0.9 | 0.92 |
| **IL-10** | 6.4 ± 2.2 | 3.0 ± 0.4 | 0.05 |
| **IL-1beta** | 18.0 ± 5.4 | 12.1 ± 3.8 | 0.69 |
| **IL-1Ra** | 17.1 ± 2.6 | 11.8 ± 0.9 | 0.68 |
| **IL-12p40** | 119.5 ± 27.0 | 67.4 ± 8.0 | 0.64 |
| **IL-13** | 138.9 ± 21.2 | 109.8 ± 11.5 | 0.99 |
| **MCP-1** | 598.4 ± 52.4 | 567.7 ± 34.3 | 0.88 |

**Supplementary Figure 2.**
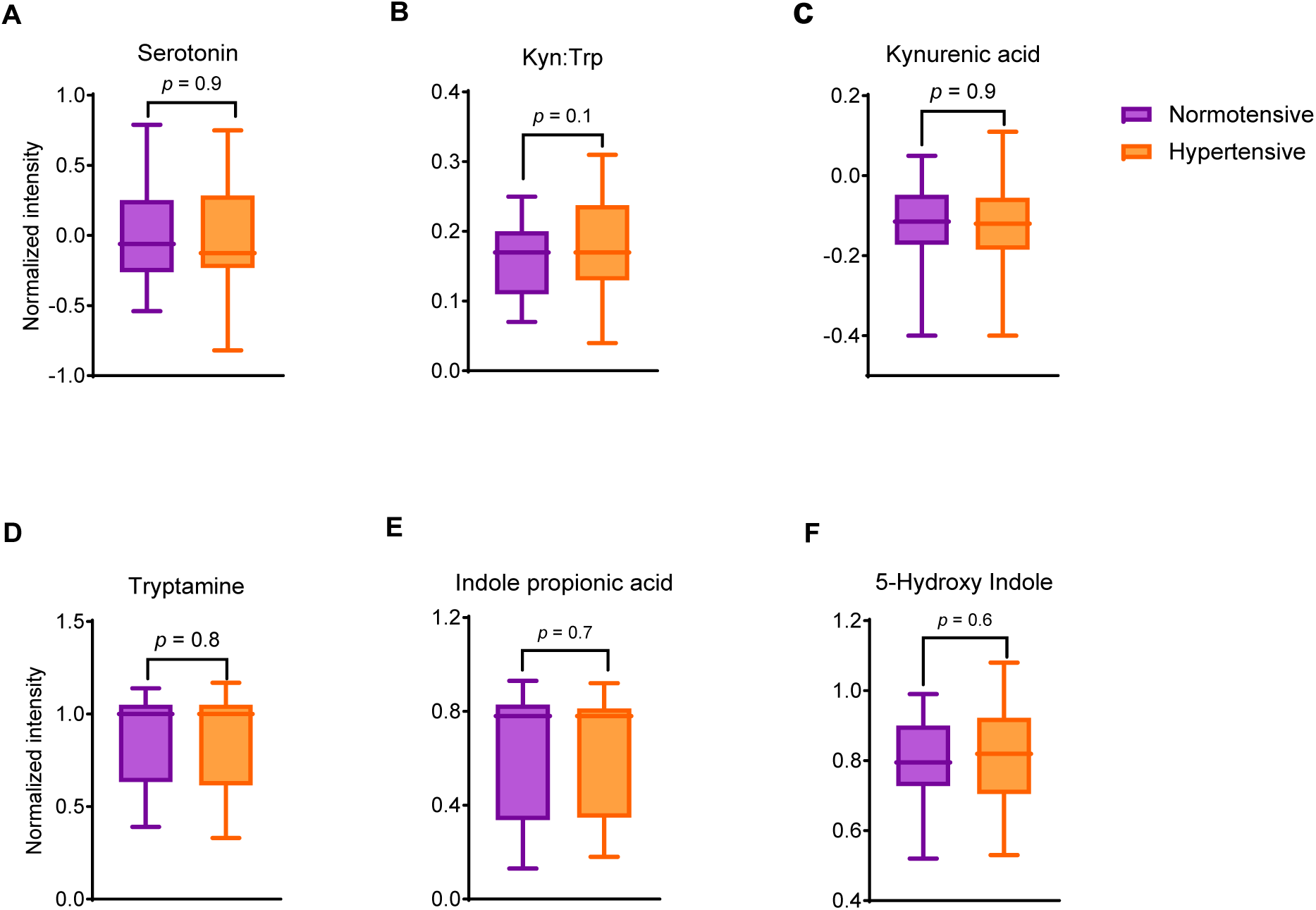
Serum concentration of tryptophan metabolites. Unpaired t-test was performed and p<0.05 was considered significant. Abbreviations: HC, healthy controls; HT, hypertensive participants.

**Supplementary Figure 3.**
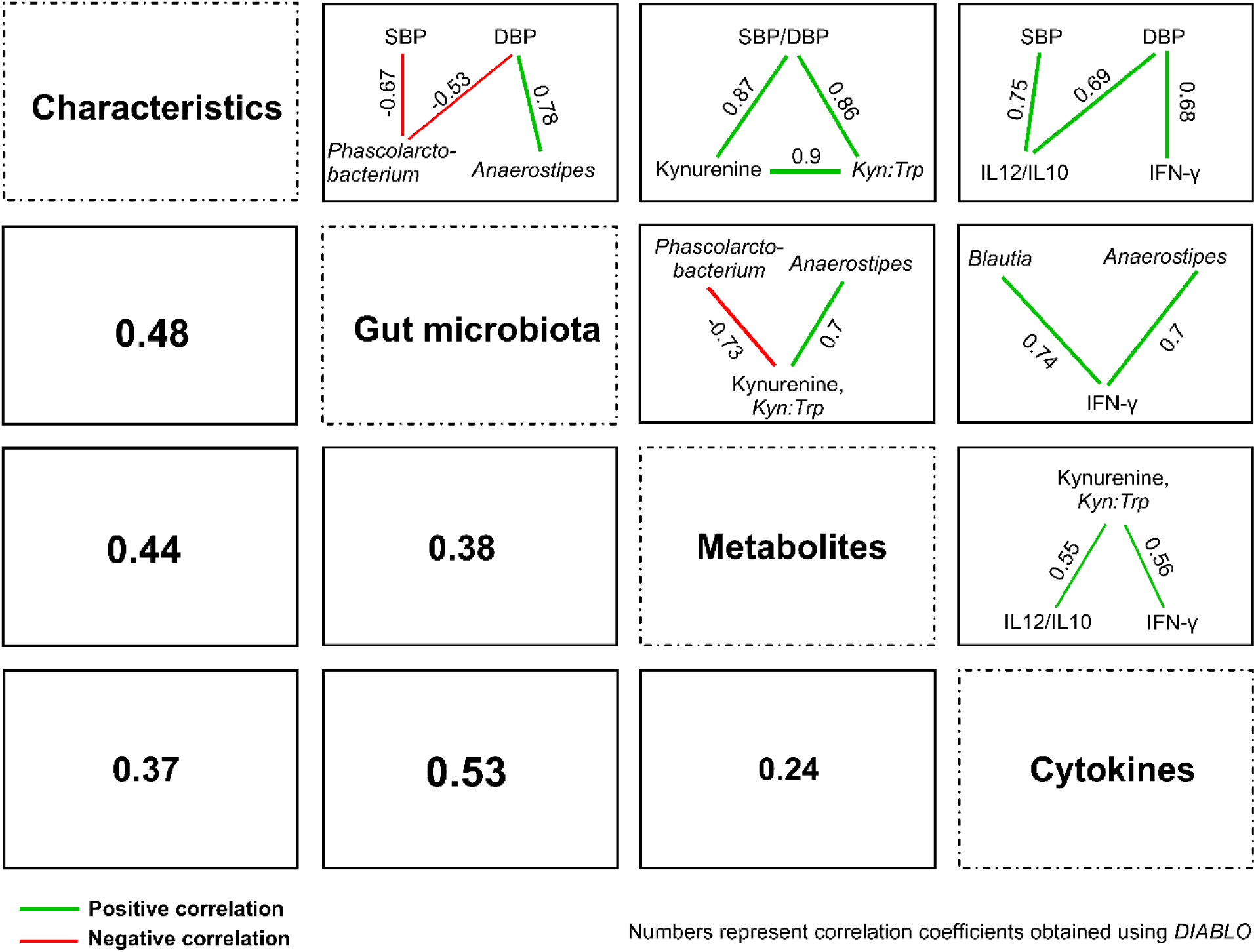
Multivariate associations. Multivariate canonical correlations between participant characteristics, microbiota, tryptophan metabolites, and cytokines datasets using **D**ata **I**ntegration **A**nalysis for **B**iomarker discovery using **L**atent c**O**mponents employing DIABLO (*mixOmics* in R). Figure illustrates most significant associations among each dataset with correlation coefficient cut-off of r > 0.55. Positive correlations are shown in green line, negative correlations are shown in red line; *r* is correlation coefficient, values closer to -1 indicates strong negative correlation and values closer to +1 indicate a strong positive correlation.

### STROBE Statement—Checklist of items that should be included in reports of *cohort studies*

|  | Item No | Recommendation | Page No. |
| --- | --- | --- | --- |
| Title and abstract | 1 | (a) Indicate the study’s design with a commonly used term in the title or the abstract | 2 |
|  |  | (b) Provide in the abstract an informative and balanced summary of what was done and what was found | 2 |
| Introduction |  |  |  |
| Background/rationale | 2 | Explain the scientific background and rationale for the investigation being reported | 5-6 |
| Objectives | 3 | State specific objectives, including any prespecified hypotheses | 6-7 |
| Methods |  |  |  |
| Study design | 4 | Present key elements of study design early in the paper | 7 |
| Setting | 5 | Describe the setting, locations, and relevant dates, including periods of recruitment, exposure, follow-up, and data collection | 7-8 |
| Participants | 6 | (a) Give the eligibility criteria, and the sources and methods of selection of participants. Describe methods of follow-up | 7 |
|  |  | (b) For matched studies, give matching criteria and number of exposed and unexposed | 7 |
| Variables | 7 | Clearly define all outcomes, exposures, predictors, potential confounders, and effect modifiers. Give diagnostic criteria, if applicable | 12-13 |
| Data sources/<br>measurement | 8 | For each variable of interest, give sources of data and details of methods of assessment (measurement). Describe comparability of assessment methods if there is more than one group | 8-10 |
| Bias | 9 | Describe any efforts to address potential sources of bias | 7, 9 |
| Study size | 10 | Explain how the study size was arrived at | 7, 41 |
| Quantitative variables | 11 | Explain how quantitative variables were handled in the analyses. If applicable, describe which groupings were chosen and why | 11-12 |
| Statistical methods | 12 | (a) Describe all statistical methods, including those used to control for confounding | 11-12 |
|  |  | (b) Describe any methods used to examine subgroups and interactions | 12 |
|  |  | (c) Explain how missing data were addressed | 9, 10 |
|  |  | (d) If applicable, explain how loss to follow-up was addressed | NA |
|  |  | (e) Describe any sensitivity analyses | 11 |
| Results |  |  |  |
| Participants | 13 | (a) Report numbers of individuals at each stage of study—eg numbers potentially eligible, examined for eligibility, confirmed eligible, included in the study, completing follow-up, and analysed | 7 |
|  |  | (b) Give reasons for non-participation at each stage | 7 |
|  |  | (c) Consider use of a flow diagram | 41 |
| Descriptive data | 14 | (a) Give characteristics of study participants (eg demographic, clinical, social) and information on exposures and potential confounders | 12-13 |
|  |  | (b) Indicate number of participants with missing data for each variable of interest | 41 |
|  |  | (c) Summarise follow-up time (eg, average and total amount) | NA |
| Outcome data | 15 | Report numbers of outcome events or summary measures over time | 13-16 |
| Main results | 16 | (a) Give unadjusted estimates and, if applicable, confounder-adjusted estimates and their precision (eg, 95% confidence interval). Make clear which confounders were adjusted for and why they were included | 11 |
|  |  | (b) Report category boundaries when continuous variables were categorized | 13 |
|  |  | (c) If relevant, consider translating estimates of relative risk into absolute risk for a meaningful time period | NA |
| Other analyses | 17 | Report other analyses done—eg analyses of subgroups and interactions, and sensitivity analyses | 11 |
| **Discussion** |  |  |  |
| Key results | 18 | Summarise key results with reference to study objectives | 16 |
| Limitations | 19 | Discuss limitations of the study, taking into account sources of potential bias or imprecision. Discuss both direction and magnitude of any potential bias | 19-20 |
| Interpretation | 20 | Give a cautious overall interpretation of results considering objectives, limitations, multiplicity of analyses, results from similar studies, and other relevant evidence | 16-19 |
| Generalisability | 21 | Discuss the generalisability (external validity) of the study results | 20 |
| **Other information** |  |  |  |
| Funding | 22 | Give the source of funding and the role of the funders for the present study and, if applicable, for the original study on which the present article is based | 23 |

